# Association of Chronotype and Shiftwork with COVID-19 Infection

**DOI:** 10.1101/2023.07.06.23292337

**Authors:** Stuart F. Quan, Matthew D. Weaver, Mark É. Czeisler, Laura K. Barger, Lauren A. Booker, Mark E. Howard, Melinda L. Jackson, Rashon I. Lane, Christine F. McDonald, Anna Ridgers, Rebecca Robbins, Prerna Varma, Shantha M.W. Rajaratnam, Charles A. Czeisler

## Abstract

**Objective:** This study assesses whether chronotype is related to COVID-19 infection and whether there is an interaction with shift work.

**Methods:** Cross-sectional survey of 19,821 U.S. adults

**Results:** COVID-19 infection occurred in 40% of participants, 32.6% morning and 17.2% evening chronotypes. After adjusting for demographic and socioeconomic factors, shift work, sleep duration and comorbidities, morning chronotype was associated with a higher (aOR: 1.15, 95% CI 1.10-1.21) and evening chronotype with a lower (aOR: 0.82, 95% CI: 0.78-0.87) prevalence of COVID-19 infection in comparison to an intermediate chronotype. Working exclusively night shifts was not associated with higher prevalence of COVID-19. Morning chronotype and working some evening shifts was associated with the highest prevalence of previous COVID-19 infection (aOR: 1.87, 95% CI: 1.28-2.74).

**Conclusion:** Morning chronotype and working a mixture of shifts increase risk of COVID-19 infection.

**Learning Outcomes:**

- Describe the association between chronotype and prevalence of COVID-19 infection
- Summarize the combined effect of chronotype and shift work on the prevalence of COVID-19 infection

## Introduction

A number of underlying medical conditions such as cardiovascular disease, diabetes, and obesity increase the risk for COVID-19 infection and severity.^1,2^ Sleep conditions also are important risk factors associated with increased prevalence and worse outcomes for COVID-19 disease; emerging evidence demonstrates that obstructive sleep apnea^3–6^ and more recently, insomnia and reduced sleep duration are linked to an increase in prevalence and worse COVID-19 disease outcomes.^7^ Circadian rhythmicity is important in the regulation of immune system and inflammatory processes.^8,9^ Chronotype is a trait that strongly reflects circadian timing as well as social and homeostatic factors.^10^ Recently, evening chronotype has been associated with greater risk of poor health with increased rates of obesity, cardiovascular disease, and diabetes.^11,12^ However, there have been few studies of whether a specific diurnal preference or chronotype is associated with a greater risk for COVID-19 disease. In a single center study in France, social jet lag as a marker of evening chronotype was associated with a higher rate of COVID-19.^13^ In contrast, the COVID-19 Outbreak Public Evaluation (COPE) Initiative reported that morning in comparison to evening chronotype was associated with a higher rate of COVID infection;^7^ while in a study using UK Biobank participants, no association was observed between a specific chronotype and COVID-19.^14^ Thus, the impact of chronotype on COVID-19 has not been determined.

During the height of the COVID-19 pandemic, sleep patterns changed. As a result of lockdowns and social restrictions, stress, meal timing, anxiety and depression, social isolation, and the migration to remote working, both increases and decreases in sleep duration as well as changes in sleep timing occurred.^15–17^ The presence of these factors may allow the development of circadian misalignment which increases risk of cardiovascular and metabolic disorders, and importantly infections. Circadian misalignment is a characteristic of shift workers who have been shown to have an increased risk of developing respiratory infections.^18,19^ It has been suggested that shift workers are at greater risk for COVID-19.^20–22^ This has been confirmed by several studies observing that night shift work is associated with higher rates of COVID-19 infection and hospitalization.^23–27^

Investigations examining whether the adverse effect of shift work on health is modified by chronotype suggest that shift workers with evening chronotype may have greater risk for diabetes and obesity.^12^ However, there have been no assessments of whether risk of infection and in particular COVID-19 is similarly affected.

In this study, we examined whether both morning and evening chronotypes were more likely to have had a COVID-19 infection in comparison to those who were intermediate chronotypes. Additionally, we examined whether there is a synergistic association between chronotype and shift work, with COVID-19 infection. To accomplish this, we used data from the first four 2022 waves of COPE Initiative (http://www.thecopeinitiative.org/), a program focused on accumulating data on public attitudes, behaviors and beliefs related to the COVID-19 pandemic from large scale, demographically representative samples.

## Methods

### Study Design and Participants

From March 10, 2022 to August 18, 2022, the COPE Initiative administered four successive waves of public health surveillance surveys. Dates of administration were: Wave 1 (March 10-30, 2022), Wave 2 (April 4-May 1, 2022), Wave 3 (May 4-June 2, 2022) and Wave 4 (July 28-August 18, 2022). Using demographic quota sampling to approximate population estimates for age, sex, race, and ethnicity based on the 2020 U.S. census, each wave consisted of more than 5000 unique participants recruited by Qualtrics, LLC (Provo, Utah, and Seattle, Washington, U.S.). The Monash University Human Research Ethics Committee (Study #24036) approved the study.

### Survey Items

Participants self-reported demographic, anthropometric, and socioeconomic information including age, race, ethnicity, sex, height and weight, education level, employment status and household income. In addition, they reported information on several current and past medical conditions by answering the question: “Have you ever been diagnosed with any of the following conditions: high blood pressure, cardiovascular disease (e.g., heart attack, stroke, angina), gastrointestinal disorder (e.g., acid reflux, ulcers, indigestion), cancer, chronic kidney disease, liver disease, sickle cell disease, chronic obstructive pulmonary disease or asthma?” Possible responses to each condition were “Never”, “Yes I have in the past, but don’t have it now”, “Yes I have, but I do not regularly take medications or receive treatment”, and “Yes I have, and I am regularly taking medications or receiving treatment”.

Each survey contained identical items related to COVID-19 infection status and the number of COVID-19 vaccinations participants had obtained. Ascertainment of past COVID-19 infection was obtained using responses from the following questions related to COVID-19 testing:

1. “Have you ever tested positive?”
2. “Despite never testing positive, are you confident that you have had COVID-19?”
3. “Despite never testing positive, have you received a clinical diagnosis of COVID-19?”
4. “Have you experienced a problem with decreased sense of smell or taste at any point since January 2020?”

Chronotype was ascertained by asking participants the following question from the Horne & Östberg Morningness-Eveningness questionnaire:”^28^ “One hears about ‘morning’ and ‘evening’ types of people. Which one of these types do you consider yourself to be?” Possible responses were “Definitely a ‘morning’ type”, “Rather more of a ‘morning’ than an ‘evening’ type”, “Rather more of an ‘evening’ than a ‘morning’ type”, “Definitely an ‘evening’ type”.^29^ Morningness was defined as definitely a “morning” type and eveningness conversely was defined as definitely an “evening” type. Rather more of an “evening” and rather more of a “morning” were classified as neither morning nor evening type (intermediate).

Sleep duration was assessed using a question from the Pittsburgh Sleep Quality Index.^30^ Responses were rounded to the nearest hour; those <3 hours or >12 hours were excluded as improbable estimates (N=987) as was done in a previous analysis of this cohort.^7^

For participants who endorsed self-employment, full or part-time employment, the following question was asked to determine the presence of shift work: “Currently, what type of shifts do you work? (Select all that apply)” Possible responses were:

1. “Day shift occurs any time between 6am and 7pm”
2. “Evening shift occurs any time between 3pm and midnight”
3. “Night shift is any shift in which the majority of the work hours occur between 10pm and 8am”

From these responses, 5 work shift categories were constructed: retired/not working, day shift only, day and/or some evening shifts, day and/or evening shifts and some night shifts, and night shift only. In addition, participants who were employed were asked to provide the percentage of their paid work hours that were completed remotely.

### Statistical Analyses

Summary data for continuous or ordinal variables are reported as their respective means and standard deviations (SD) and for categorical variables as their percentages. Consistent with previous analyses, we defined a positive history of COVID-19 infection as an affirmative response to having tested positive for COVID-19, new loss of taste or smell or a clinical diagnosis of COVID-19. Number of COVID-19 vaccinations was utilized as an ordinal variable and also dichotomized as boosted (>2 vaccinations) or not boosted (≤2 vaccinations). Comorbid medical conditions were defined as currently having the condition whether treated or untreated. The effect of comorbid medical conditions was evaluated by summing the number of conditions reported by the participant (minimum value 0, maximum value 9). Body mass index (BMI) was calculated using self-reported height and weight as kg/m^2^. Socioeconomic covariates were dichotomized as follows: employment (retired vs. not retired), education (high school or less vs. some college or higher) and income in U.S. Dollars to approximate 200% of the 2022 U.S. Poverty Level for a family of 4 (<$50,000 vs ≥$50,000).^31^

Comparisons of continuous or ordinal variables stratified by COVID-19 infection status were performed using Student’s unpaired t-test. Bivariate comparisons of categorical variables stratified by COVID-19 infection were completed using χ^2^.

Multivariable modelling using logistic regression was utilized to determine whether circadian preference and shift work categories were associated with COVID-19 infection. In an initial analysis including all participants with complete data, an initial baseline model was constructed only entering chronotype as a categorical variable with morningness and eveningness compared to the referent of neither morningness nor eveningness. We then developed increasingly complex models by sequentially including demographic factors, comorbidities and vaccination status, socioeconomic factors, shift work category, and sleep duration. In a subsequent model, to separately determine the joint effects of diurnal preference and shift work on the prevalence of COVID-19 infection, analysis was limited only to participants who were full-time, part-time, or self-employed. Models sequentially included demographic factors, comorbidities and vaccination status, socioeconomic factors, and sleep duration as well as 2-way interactions between diurnal preference and shift work categories. In addition, we performed sensitivity analyses with stricter (i.e., using COVID-19 infection as a positive test only) and broader (i.e., our original definition plus presumed positive, but not tested as an indicator of a past COVID-19 infection) definitions.

All analyses were conducted using IBM SPSS version 28 (Armonk, NY). A p<0.05 was considered statistically significant.

## Results

Table 1 shows the bivariate associations between COVID-19 infection status, diurnal preference, and co-morbid medical, demographic, and social characteristics of the cohort. Of the 19821 participants, 7932 (40.0%) had at least one COVID-19 infection. Morning and evening diurnal preferences were observed in 31.9% and 19.4% of participants respectively. Among COVID-19 positive in comparison to COVID-19 negative participants, there was a higher percentage of morning types (32.6 vs 31.4%) and a lower percentage of evening types (17.2 vs. 20.9%, p≤0.001). Participants who were retired or unemployed were less likely to have had a COVID-19 infection (12.5 vs. 30.8%, p≤0.001). COVID-19 positive in comparison to COVID-19 negative participants were younger (40.2 ± 15.8 vs. 50.4 ± 18.0 y, p≤0.001), had a slightly shorter self-reported sleep duration (6.8 ± 1.6 vs. 6.9 ± 1.7 h, p≤0.001), had a slightly larger percent of time working remotely (36 ± 34 vs. 32 ± 38%, p≤0.001), were slightly more overweight (BMI: 28.7 vs. 28.3 kg/m^2^, p<0.001) had more comorbidities (1.7 ± 2.4 vs. 0.8 ± 1.2, p≤0.001), and were less likely to have received a COVID-19 booster vaccination (27.6 vs. 46.4%, p≤0.001). COVID-19 positive participants also were more likely to be Hispanic, not retired and have an income more than $50,000 per year.

**Table 1:**
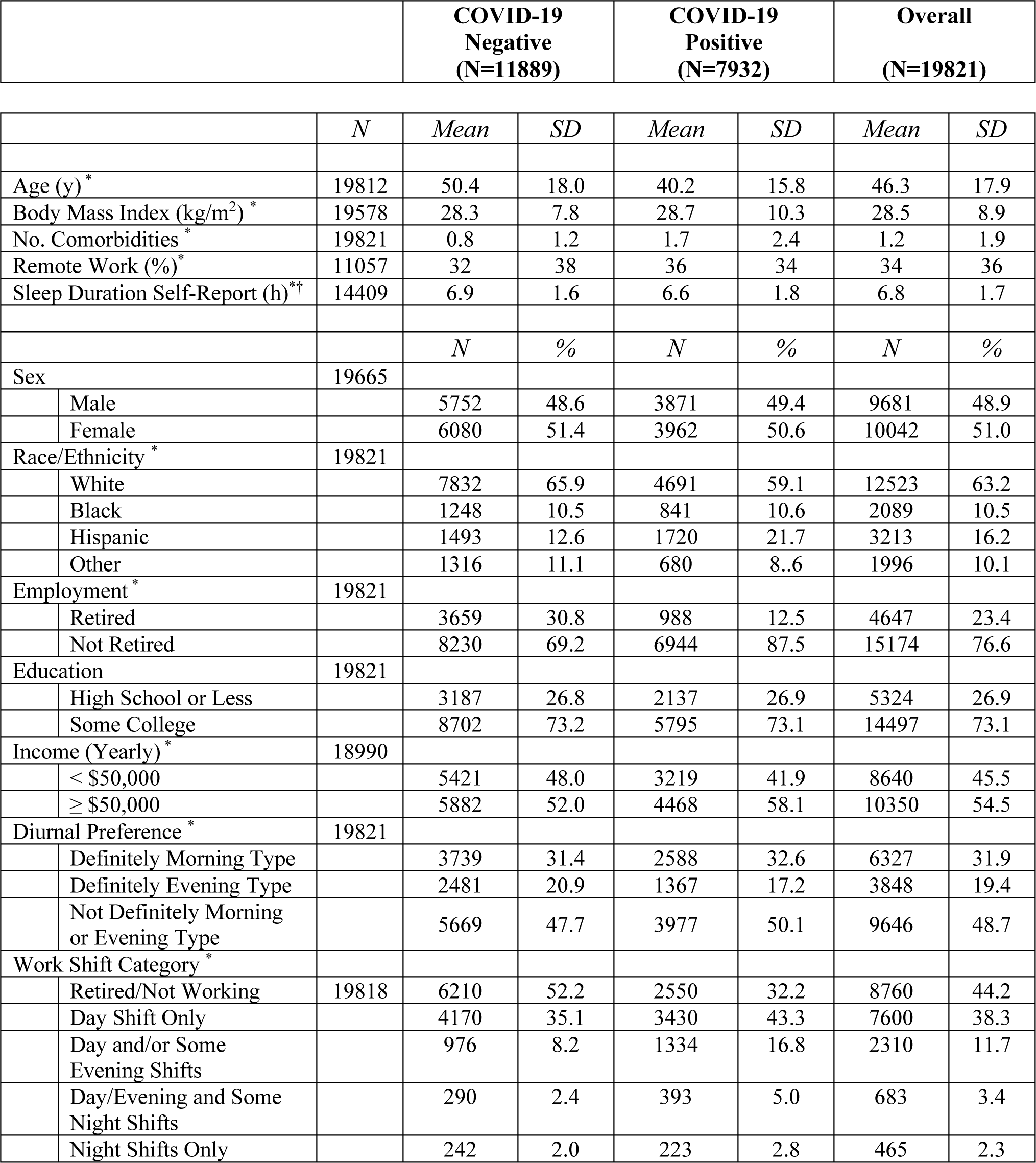
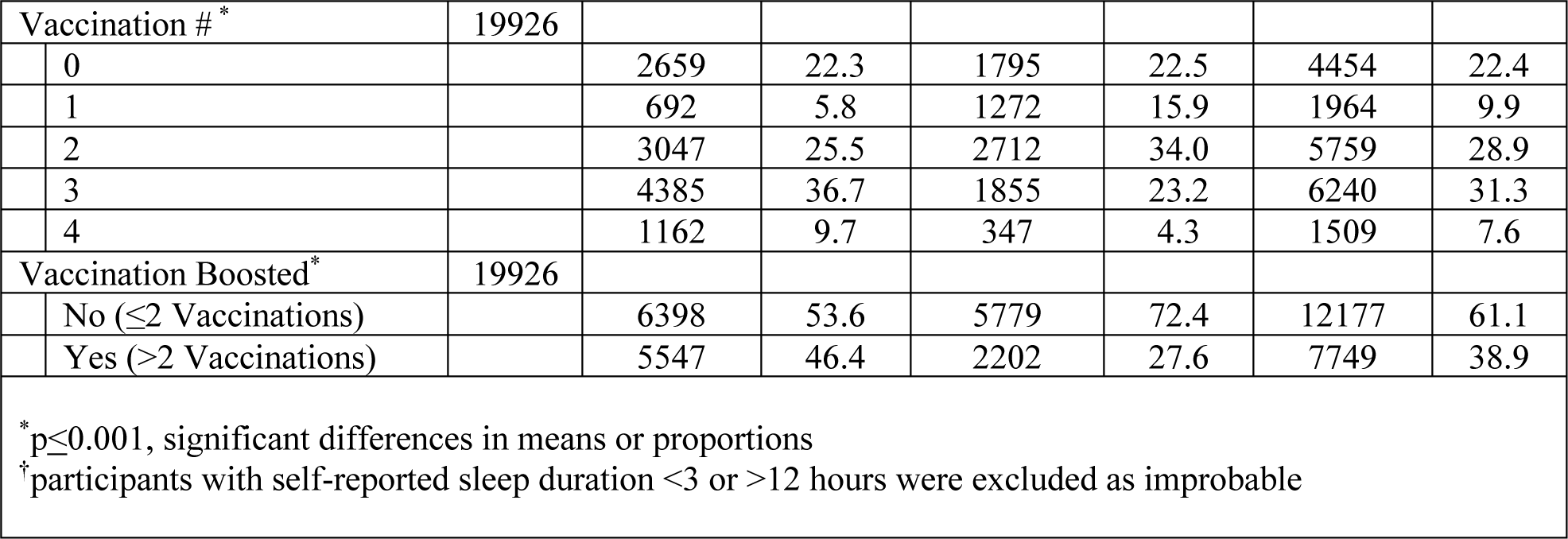
Associations Between COVID-19 Infection Status, Diurnal Preference, Shift Work, and Co-morbid Medical, Demographic and Social Characteristics

Table 2 displays the logistic regression models for the association between having had a COVID-19 infection and either a morning or evening diurnal preference in comparison to a reference group without a definite preference. In models including demographics, comorbidities, vaccination status, and socioeconomic factors, morning preference was related to a 20% greater likelihood of COVID-19 infection. In contrast, evening preference was 20% less likely to be linked to a COVID-19 infection. These associations were not materially affected by the type of work shift, percent time working remotely or after adjustment for sleep duration.

**Table 2:** Odds Ratio (adjusted) for Reporting One or More COVID-19 Infections Based on Circadian Preference (N=19821)

| *Model* | **Morningness** |  | **Eveningness** |  |
| --- | --- | --- | --- | --- |
|  | *aOR* | *95% CI* | *aOR* | *95% CI* |
| Baseline | 1.07 | 1.03-1.12* | 0.86 | 0.81-0.90* |
| +Demographics<sup>†</sup> | 1.21 | 1.15-1.26* | 0.76 | 0.73-0.81* |
| +Comorbidities and Vaccination Status<sup>‡</sup> | 1.23 | 1.16-1.31* | 0.77 | 0.72-0.83* |
| +Socioeconomic<sup>§</sup> | 1.20 | 1.13-1.28* | 0.80 | 0.74-0.86* |
| +Shift Work | 1.21 | 1.14-1.29* | 0.79 | 0.73-0.85* |
| +Self-Reported Sleep Duration and % Remote Work | 1.23 | 1.15-1.31* | 0.78 | 0.73-0.85* |

The baseline model includes only definitely morning or definitely evening preference. Subsequent models are additive to their immediate predecessor and are adjusted as indicated below (see text for covariate definitions) with the fully adjusted model reflecting demographic, comorbid disease, socioeconomic characteristics, and the occurrence of shift work.
<sup>†</sup>age, sex, race
<sup>‡</sup>BMI, vaccination status (boosted vs. not boosted), # of the following conditions: diabetes, asthma, sickle cell disease, cardiovascular disease, hypertension, cancer, chronic kidney disease, liver disease, chronic obstructive pulmonary disease
<sup>§</sup>education, income, employment
\*p<0.001

In Table 3 are the bivariate associations between COVID-19 infection and diurnal preference, and co-morbid medical, demographic, and social characteristics of the cohort limited only to members who were employed. In comparison to the entire cohort, participants were slightly younger, more likely to be male, and had a higher income. Participants who worked a mixture of shift types had a higher prevalence rate of infection in comparison to those who worked only day shifts or only night shifts (p<0.001).

**Table 3:**
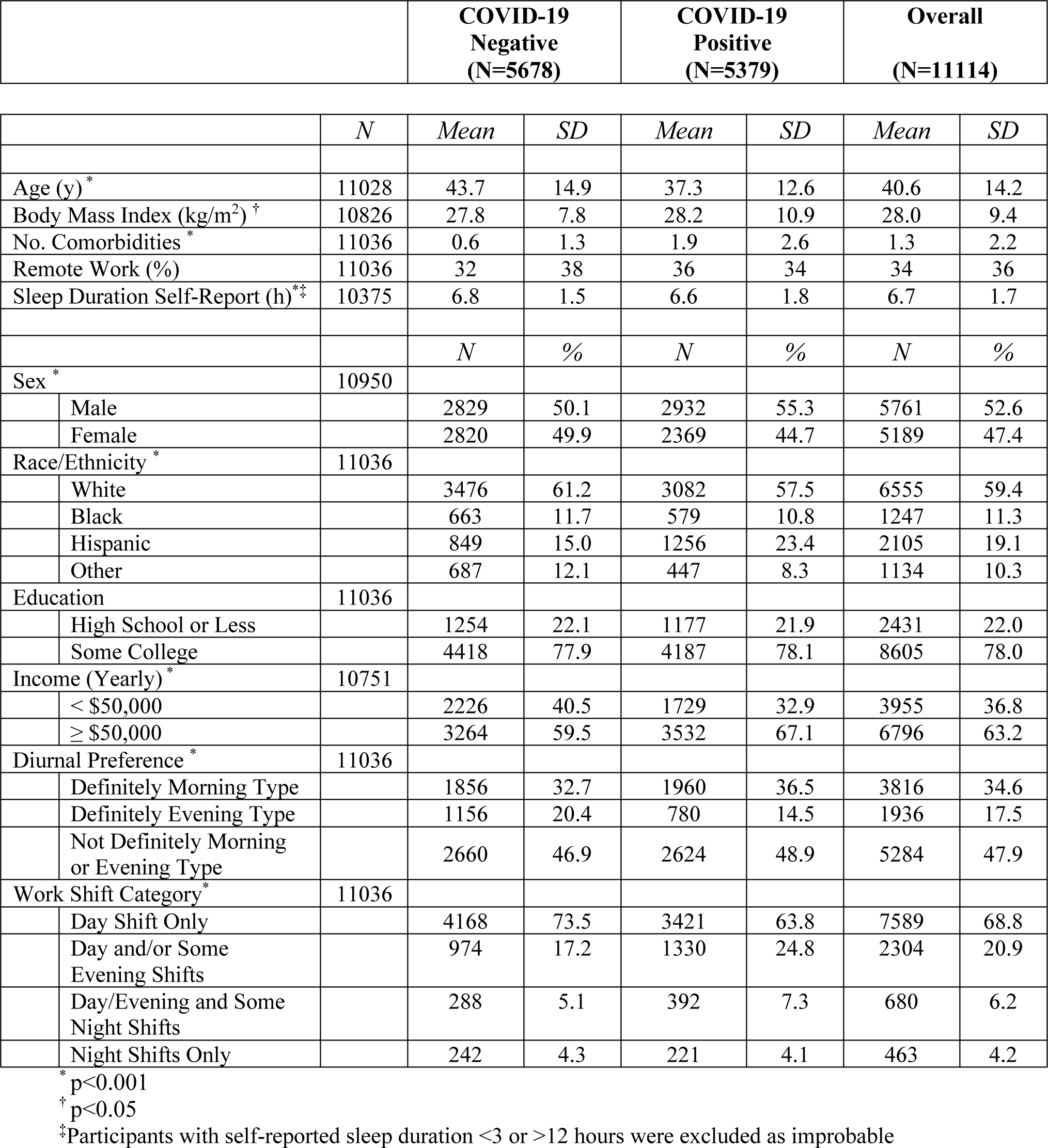
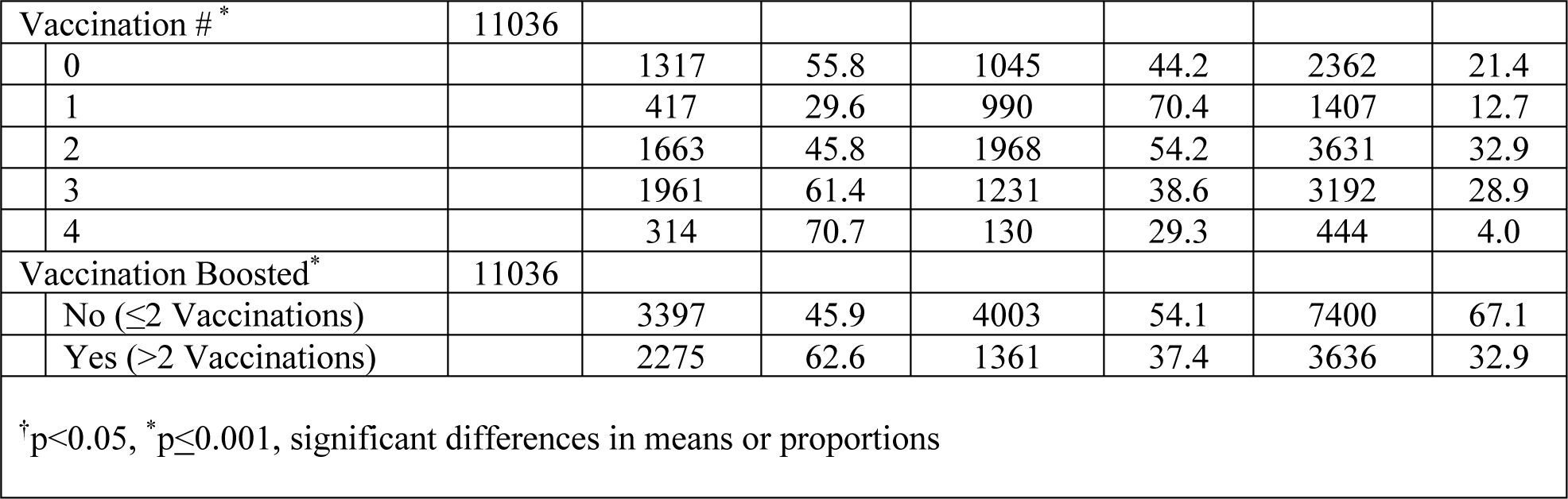
Associations Between COVID-19 Infection Status, Diurnal Preference, Shift Work, and Co-morbid Medical, Demographic and Social Characteristics in Employed Participants

Table 4 shows the fully adjusted logistic regression model which adjusted for percent remote work and sleep duration documenting the association between COVID-19 infection and diurnal preference, work shift category and the interactions between diurnal preference and work shift category. In comparison to neither definite morning nor evening preference, there was a 18% greater likelihood of infection in those with morning preference and a 16% lower likelihood in those with an evening preference. Both were comparable to the aORs observed for the overall cohort. In comparison to working only a day shift, a greater risk of COVID-19 infection was observed in those working a mixture of day and evening shifts (aOR: 1.25, 95% CI: 1.10-1.41); working a mixture of day, evening, and night shifts approached significance (aOR: 1.18, 95% CI: 0.97-1.44). Working only night shifts was not associated with a higher prevalence of COVID-19 infection. The overall interaction between diurnal preference and work shift category was significant (Wald χ^2^=13.864, p=0.031). Individual interaction terms indicated that there was a higher likelihood of infection in participants who had a morning preference and working a combination of day and evening shifts (aOR: 1.87, 95% CI: 1.28-2.74); the association with those with a morning preference and working a mixture of day, evening, and night shifts approached significance (aOR: 1.72, 95% CI:0.98-3.02).

**Table 4:** Odds Ratio (adjusted) in Fully Adjusted Model* with Interactions for Reporting One or More COVID-19 Infections Based on Circadian Preference and Shift Work (N=9877)

|  | **aOR** | **95%CI** |
| --- | --- | --- |
| **Diurnal Preference** |  |  |
| Neither Morning nor Evening Preference | Referent |  |
| Definite Morning Preference | 1.18 | 1.09-1.27 |
| Definite Evening Preference | 0.84 | 0.76-0.92 |
| **Work Shift Category** |  |  |
| Day Shift Only | Referent |  |
| Day and/or Some Evening Shifts | 1.25 | 1.10-1.41 |
| Day/Evening and Some Night Shifts | 1.18 | 0.97-1.44 |
| Night Shifts Only | 0.95 | 0.75-1.20 |
| **Diurnal Preference x Work Shift Interactions** |  |  |
| Neither Morning nor Evening Preference x Day Shift Only | Referent |  |
| Morning Preference x Day and/or Some Evening Shifts | 1.87 | 1.28-2.74 |
| Morning Preference x Day/Evening and Some Night Shifts | 1.72 | 0.98-3.02 |
| Morning Preference x Night Shifts Only | 0.93 | 0.47-1.84 |
| Evening Preference x Day and/or Some Evening Shifts | 0.89 | 0.59-1.33 |
| Evening Preference x Day/Evening and Some Night Shifts | 0.68 | 0.39-1.24 |
| Evening Preference x Night Shifts Only | 0.79 | 0.41-1.52 |

\*Fully adjusted model includes age, sex, race, BMI, vaccination status (boosted vs. not boosted), # of the following conditions: diabetes, asthma, sickle cell disease, cardiovascular disease, hypertension, cancer, chronic kidney disease, liver disease, chronic obstructive pulmonary disease, education, income, employment, sleep duration, percent time working remotely

Sensitivity analyses indicated that our findings were similar using slightly stricter or more liberal definitions of COVID-19. However, a definition that required a positive test only lost statistical significance.

## Discussion

In this study, morning and evening chronotypes were found to be differentially associated with COVID-19 infection; morning types were more likely and evening types were significantly less likely to be infected. Additionally, in comparison to working only day shifts, working a combination of different shift types, as opposed to day shifts only was associated with a higher prevalence of COVID-19 infection. This finding of higher infection rate was primarily observed in individuals with a morning chronotype who worked a combination of different shifts.

Morning chronotypes were more susceptible to COVID-19 infection than evening chronotypes; this finding is consistent with earlier preliminary results from the same cohort.^7^ This observation stands in contrast to a large body of evidence demonstrating that the prevalence of cardiovascular disease risk factors and metabolic disorders is higher in evening chronotypes.^11,12^ Furthermore, our results are distinct from the limited studies that also have assessed the association between chronotype and COVID-19. In a single center study, social jet lag as a marker of evening chronotype was found to be associated with a higher rate of COVID-19.^13^ In contrast, no association between chronotype and COVID-19 infection was observed in two separate analyses of the UK Biobank^14,26^ as well as in an analysis of long COVID from the Nurses’ Health Study.^32^ Additionally, to our knowledge, there have been no previous studies of an association between chronotype and infection risk from other pathogens.

Despite the paucity of clinical investigations related to chronotype and infection risk, there are mechanisms that could explain our finding of an association between morning chronotype and COVID-19 infection. A reduction in sleep duration has been linked to increased susceptibility to infection.^19^ However, our observation remained robust after controlling for sleep duration in our modelling. Alternatively, various components of the immune system exhibit circadian rhythmicity.^33^ Secretion of inflammatory cytokines, trafficking myeloid and lymphocyte subsets and maturation of leukocytes exhibit a circadian rhythm with the aggregate response favoring a proinflammatory response while asleep.^33^ Some,^34,35^ but not all^36,37^ have found that time of day influences the antibody response to COVID-19 vaccination. Therefore, it is possible that immunity to COVID-19 is less robust in morning chronotypes resulting in greater susceptibility to infection.

In this study, the association between working night shift and COVID-19 infection was the same as working day shift. However, working a mixture of shifts, particularly ones that included evening shifts was associated with a higher likelihood of COVID-19 infection. These findings stand in contrast to some previous observations that have noted a positive association between rotating and consistent/permanent night shift work and COVID-19 infection.^24,25,27^ However, not all studies have found a significant relationship among those who only worked consistent/permanent night shift.^23,26^ Working night shifts and early morning shifts are associated with circadian misalignment as well as shorter sleep duration. Both may increase the propensity for COVID-19 infection as well as other respiratory contagions. However, night shift workers tend to have less interpersonal contact than day shift workers.^38^ This may mitigate the adverse impact of circadian misalignment on increased risk of COVID-19 infection. Our results suggest that the impact of circadian misalignment and sleep loss, and consequent predisposition towards being infected with COVID-19 is greater among persons who irregularly perform shift work rather than in those who work permanent/consistent day or night shifts.

We observed a significant interaction between chronotype and shift work in which morning diurnal preference together with working a mixture of shifts was associated with the highest aOR of having had a COVID-19 infection. This implies that irregularity in sleep schedule was a major factor in susceptibility to COVID-19 infection in those with morning diurnal preference. Irregularity in sleep-wake patterns has been associated with higher levels of cardiovascular disease biomarkers^39^ as well as a greater risk of cardiovascular disease.^40^ Using UK Biobank data, sleep irregularity also has been demonstrated to increase risk of COVID-19 infection and its severity.^41^ Potential explanatory mechanisms include the impact of sleep irregularity on reducing sleep duration as well as consequences of higher levels of circadian misalignment. Both of these factors can contribute to greater amounts of both acute and chronic inflammation resulting in higher risk of COVID-19 infection.

Our study should be interpreted in the context of several limitations. First, all of the data were self-reported including ascertainment of both diurnal preference (using a single self-report item) and COVID-19 infection. However, sensitivity analyses using different definitions of COVID-19 infection were qualitatively similar to the findings reported herein. In addition, broadly similar to a previous report, we found that morning chronotypes were more prevalent than evening chronotypes, suggesting that our ascertainment of chronotype was acceptable.^42^ Second, our analyses were cross-sectional and causal inference cannot be assumed. Third, although we attempted to adjust for a number of factors known to increase risk of COVID-19 infection, residual confounding is possible.

In conclusion, morning diurnal preference is associated with an increase and evening diurnal preference is associated with a decrease in COVID-19 infection. Neither working day nor night shifts were linked to an increase in prevalence of COVID-19 infection. However, COVID-19 infection was more likely in those working a mixture of shifts, with the greatest risk conferred on those with a morning chronotype. These findings may be informative for developing measures for greater COVID-19 surveillance of shift workers with a morning chronotype, particularly as first responders in healthcare and other settings are often engaged in shift work.

## Data Availability

All data produced in the present study are available upon reasonable request to the authors

## Acknowledgments

Concept and design: SFQ

Data collection: MDW, MÉC, MEH

Data analysis and interpretation: SFQ, LAB, JFW, PV, RR, MÉC Drafting of the manuscript: SFQ

Critical feedback and revision of the manuscript: MDW, MÉC, LKB, LAB, MEH, MLJ, RL, CFM, AR, RR, PV, SMWR, CAC

## References

1. Garg S. Hospitalization Rates and Characteristics of Patients Hospitalized with Laboratory-Confirmed Coronavirus Disease 2019 — COVID-NET, 14 States, March 1–30, 2020. MMWR Morb Mortal Wkly Rep. 2020;69:458–464.

2. Ioannou GN, Locke E, Green P, et al. Risk Factors for Hospitalization, Mechanical Ventilation, or Death Among 10 131 US Veterans With SARS-CoV-2 Infection. JAMA Netw Open. 2020;3:e2022310.

3. Cade BE, Dashti HS, Hassan SM, Redline S, Karlson EW. Sleep Apnea and COVID-19 Mortality and Hospitalization. Am J Respir Crit Care Med. 2020;202:1462–1464.

4. Chung F, Waseem R, Pham C, et al. The association between high risk of sleep apnea, comorbidities, and risk of COVID-19: a population-based international harmonized study. Sleep Breath Schlaf Atm. 2021;25:849–860.

5. Maas MB, Kim M, Malkani RG, Abbott SM, Zee PC. Obstructive Sleep Apnea and Risk of COVID-19 Infection, Hospitalization and Respiratory Failure. Sleep Breath Schlaf Atm. 2021;25:1155–1157.

6. Mashaqi S, Lee-Iannotti J, Rangan P, et al. Obstructive sleep apnea and COVID-19 clinical outcomes during hospitalization: a cohort study. J Clin Sleep Med JCSM Off Publ Am Acad Sleep Med. 2021;17:2197–2204.

7. Quan SF, Weaver MD, Czeisler MÉ, et al. Insomnia, Poor Sleep Quality and Sleep Duration and Risk for COVID-19 Infection and Hospitalization. Am J Med. 2023;S0002-9343(23)00248–6.

8. Jerigova V, Zeman M, Okuliarova M. Circadian Disruption and Consequences on Innate Immunity and Inflammatory Response. Int J Mol Sci. 2022;23:13722.

9. Wang C, Lutes LK, Barnoud C, Scheiermann C. The circadian immune system. Sci Immunol. 2022;7:eabm2465.

10. Raman S, Coogan AN. Chapter 47 - Closing the Loop Between Circadian Rhythms, Sleep, and Attention Deficit Hyperactivity Disorder. In: Dringenberg HC, editor. Handbook of Behavioral Neuroscience. Elsevier. p. 707–716.

11. Lotti S, Pagliai G, Colombini B, Sofi F, Dinu M. Chronotype Differences in Energy Intake, Cardiometabolic Risk Parameters, Cancer, and Depression: A Systematic Review with Meta-Analysis of Observational Studies. Adv Nutr Bethesda Md. 2022;13:269–281.

12. Hittle BM, Gillespie GL. Identifying shift worker chronotype: implications for health. Ind Health. 2018;56:512–523.

13. Coelho J, Micoulaud-Franchi J-A, Wiet A-S, Nguyen D, Taillard J, Philip P. Circadian misalignment is associated with Covid-19 infection. Sleep Med. 2022;93:71–74.

14. Liu Z, Luo Y, Su Y, et al. Associations of sleep and circadian phenotypes with COVID-19 susceptibility and hospitalization: an observational cohort study based on the UK Biobank and a two-sample Mendelian randomization study. Sleep. 2022;45:zsac003.

15. Batool-Anwar S, Robbins R, Ali SH, et al. Examining Changes in Sleep Duration Associated with the Onset of the COVID-19 Pandemic: Who is Sleeping and Who is Not? Behav Med Wash DC. 2021;1–10.

16. Wright KP, Linton SK, Withrow D, et al. Sleep in university students prior to and during COVID-19 Stay-at-Home orders. Curr Biol CB. 2020;30:R797–R798.

17. Robbins R, Affouf M, Weaver MD, et al. Estimated Sleep Duration Before and During the COVID-19 Pandemic in Major Metropolitan Areas on Different Continents: Observational Study of Smartphone App Data. J Med Internet Res. 2021;23:e20546.

18. Mohren DCL, Jansen NWH, Kant Ij, Galama JMD, van den Brandt PA, Swaen GMH. Prevalence of Common Infections Among Employees in Different Work Schedules: J Occup Environ Med. 2002;44:1003–1011.

19. Prather AA, Carroll JE. Associations between sleep duration, shift work, and infectious illness in the United States: Data from the National Health Interview Survey. Sleep Health. 2021;7:638–643.

20. Silva FR da, Guerreiro R de C, Andrade H de A, Stieler E, Silva A, de Mello MT. Does the compromised sleep and circadian disruption of night and shiftworkers make them highly vulnerable to 2019 coronavirus disease (COVID-19)? Chronobiol Int. 2020;37:607–617.

21. Lim RK, Wambier CG, Goren A. Are night shift workers at an increased risk for COVID-19? Med Hypotheses. 2020;144:110147.

22. Gao L, Li P, Lane JM. Sleep and circadian phenotypes: risk factors for COVID-19 severity? Sleep. 2022;45:zsac116.

23. Bjorvatn B, Merikanto I, Reis C, et al. Shift workers are at increased risk of severe COVID-19 compared with day workers: Results from the international COVID sleep study (ICOSS) of 7141 workers. Chronobiol Int. 2023;40:114–122.

24. Fatima Y, Bucks RS, Mamun AA, et al. Shift work is associated with increased risk of COVID-19: Findings from the UK Biobank cohort. J Sleep Res.;30. Epub ahead of print October 2021. DOI: 10.1111/jsr.13326.

25. Loef B, Dollé MET, Proper KI, van Baarle D, Initiative LCR, van Kerkhof LW. Night-shift work is associated with increased susceptibility to SARS-CoV-2 infection. Chronobiol Int. 2022;39:1100–1109.

26. Maidstone R, Anderson SG, Ray DW, Rutter MK, Durrington HJ, Blaikley JF. Shift work is associated with positive COVID-19 status in hospitalised patients. Thorax. 2021;76:601– 606.

27. Rizza S, Coppeta L, Grelli S, et al. High body mass index and night shift work are associated with COVID-19 in health care workers. J Endocrinol Invest. 2021;44:1097– 1101.

28. Horne J, Östberg, O. A self-assessment questionnaire to determine morningness-eveningness in human circadian rhythms. Int J Chronobiol. 1976;4:97–110.

29. Turco M, Corrias M, Chiaromanni F, et al. The self-morningness/eveningness (Self-ME): An extremely concise and totally subjective assessment of diurnal preference. Chronobiol Int. 2015;32:1192–1200.

30. Buysse DJ, Reynolds CF, Monk TH, Berman SR, Kupfer DJ. The Pittsburgh Sleep Quality Index: a new instrument for psychiatric practice and research. Psychiatry Res. 1989;28:193–213.

31. Federal Poverty Level (FPL) - Glossary. HealthCare.gov Available from: https://www.healthcare.gov/glossary/federal-poverty-level-fpl. Accessed February 13, 2023.

32. Wang S, Huang T, Weisskopf MG, Kang JH, Chavarro JE, Roberts AL. Multidimensional Sleep Health Prior to SARS-CoV-2 Infection and Risk of Post–COVID-19 Condition. JAMA Netw Open. 2023;6:e2315885.

33. Schmitz NCM, van der Werf YD, Lammers-van der Holst HM. The Importance of Sleep and Circadian Rhythms for Vaccination Success and Susceptibility to Viral Infections. Clocks Sleep. 2022;4:66–79.

34. Wang W, Balfe P, Eyre DW, et al. Time of Day of Vaccination Affects SARS-CoV-2 Antibody Responses in an Observational Study of Health Care Workers. J Biol Rhythms. 2022;37:124–129.

35. Zhang H, Liu Y, Liu D, et al. Time of day influences immune response to an inactivated vaccine against SARS-CoV-2. Cell Res. 2021;31:1215–1217.

36. Matryba P, Gawalski K, Ciesielska I, et al. The Influence of Time of Day of Vaccination with BNT162b2 on the Adverse Drug Reactions and Efficacy of Humoral Response against SARS-CoV-2 in an Observational Study of Young Adults. Vaccines. 2022;10:443.

37. Yamanaka Y, Yokota I, Yasumoto A, Morishita E, Horiuchi H. Time of Day of Vaccination Does Not Associate With SARS-CoV-2 Antibody Titer Following First Dose of mRNA COVID-19 Vaccine. J Biol Rhythms. 2022;37:700–706.

38. Cheng P, Drake CL. Psychological Impact of Shift Work. Curr Sleep Med Rep. 2018;4:104– 109.

39. Full KM, Huang T, Shah NA, et al. Sleep Irregularity and Subclinical Markers of Cardiovascular Disease: The Multi-Ethnic Study of Atherosclerosis. J Am Heart Assoc. 2023;12:e027361.

40. Huang T, Mariani S, Redline S. Sleep Irregularity and Risk of Cardiovascular Events: The Multi-Ethnic Study of Atherosclerosis. J Am Coll Cardiol. 2020;75:991–999.

41. Rowlands AV, Kloecker DE, Chudasama Y, et al. Association of Timing and Balance of Physical Activity and Rest/Sleep With Risk of COVID-19: A UK Biobank Study. Mayo Clin Proc. 2021;96:156–164.

42. Patterson F, Malone SK, Lozano A, Grandner MA, Hanlon AL. Smoking, Screen-Based Sedentary Behavior, and Diet Associated with Habitual Sleep Duration and Chronotype: Data from the UK Biobank. Ann Behav Med Publ Soc Behav Med. 2016;50:715–726.

